# Impact Of Adaptive Natural Killer Cells, KLRC2 Genotype and Cytomegalovirus Reactivation On Late Mortality In Patients With Severe Covid-19 Lung Disease

**DOI:** 10.1101/2021.10.11.21264805

**Authors:** Sarita Rani Jaiswal, Jaganath Arunachalam, Ashutosh Bhardwaj, Ashraf Saifullah, Rohit Lakhchaura, Mayank Soni, Gitali Bhagawati, Suparno Chakrabarti

## Abstract

**Objective:** COVID-19 infection results in severe lung disease in a small but significant number of infected patients. The etiopathogenesis in a subset of such patients, who continue to have progressive pulmonary disease following virus clearance remains unexplored.

**Methods:** We investigated the role of NKG2C^+^/NKG2A^-^ adaptive natural killer (ANK) cells, KLRC2 genotype and cytomegalovirus reactivation in 22 such patients.

**Results:** The median duration of virus positivity was 23 days and the median duration of hospitalisation was 48 days. The overall survival at 60 days in this group was 50%. Older age and comorbidities impacted survival negatively. CMV viremia was documented in 11 patients, with a survival of 25% vs 80% in those without viremia with viral load correlating with mortality. ANK cells were markedly depressed in all patients at day 15. However, persistently low ANK cells at 30 days along with an inversely high NKG2C^-^/NKG2A^+^ inhibitory NK cells significantly correlated with high CMV viremia as well as mortality, irrespective of KLRC2 genotype. Day 30 ANK cells were significantly lower in KLRC2 deletion group. IFN-gamma and Perforin release were severely compromised in all patients at day +15, with significant improvement in the survivors at day +30, but not in those with adverse outcome.

**Conclusion:** Patients with severe lung disease even after negative SARS-CoV-2 status, with persistently reduced and functionally compromised ANK cells, are more likely to have CMV reactivation and an adverse outcome, independent of KLRC2 genotype.

## Introduction

The recent pandemic caused by SARS-CoV-2 has been marked by morbidity and mortality due to lung disease in a small but significant number of infected patients(1). Disease progression and adverse outcomes have been observed even after viral clearance in some(2). This raises the question about the etiopathogenesis of progressive disease and late death even after decrease in virus load.

Natural killer (NK) cells have traditionally been considered as front-line of defence against viral pathogens. However, the canonical NK cells are not antigen specific and have a short lifespan. On the other hand, long-lasting NK cells with virus-specific memory following exposure to cytomegalovirus (CMV) in both mice as well as humans, have gained attention in the last decade(3). These cells, now commonly referred to as adaptive NK (ANK) cells, are characterised by strong expression of NKG2C, which is an activating receptor with HLA-E as it’s ligand. NKG2A, which provides a strong inhibitory signal to NK cells on binding to the same ligand, is not expressed on these cells, enabling them with interferon-gamma (IFN-g) release potential as well as strong cytotoxicity(4).

At the early stages of COVID-19 pandemic, our group had hypothesised that ANK cells might have a significant impact on the severity and outcome of infection with SARS-CoV-2(5). This was based on our observations of strong correlation between an anti-leukemia as well as an antiviral effect of ANK cells derived from CMV-seropositive donors(6-8). Subsequently, a few studies have studied the NK cell profile in COVID-19 patients during the acute phase(9). Deletion of KLRC2 gene, which encodes for NKG2C, has been described as a risk factor for severe disease(10). However, longitudinal study of ANK cells and their impact on late complications in those with prolonged COVID-19 disease is lacking.

During the recent surge of COVID-19 infection in India witnessed between April-June 2021, we prospectively evaluated non-immunocompromised patients with severe lung-disease surviving beyond 15 days from diagnosis, for KLRC2 genotype, ANK cells and CMV viremia, with respect to their long-term outcome.

### Patients and Methods

Patients with severe COVID-19 related lung disease diagnosed between 1^st^ April to 15^th^ May 2021, who survived for more than 15 days from the onset of the illness were followed up with weekly RT-PCR for SARS-CoV-2. Patients with malignant diseases and chronic renal failure were excluded. They were evaluated on days 15 and 30 of their illness for CMV reactivation and immunological parameters as listed below. The study was approved by institutional ethics committee.

### Treatment for COVID-19 lung disease

All patients received intravenous methylprednisolone (2 mg/kg) for 3 days, followed by tapering over next 7 days, remdesivir, low molecular weight heparin and broad-spectrum antibiotics. All required intensive care admission and invasive or non-invasive ventilation support.

### Real Time Reverse Transcription Polymerase chain (RT-PCR) for Detection of SARS-CoV-2

All Covid tests were done by Truenat Real Time Reverse transcription Polymerase chain (RT-PCR) test. Samples taken were Oropharyngeal or Nasopharyngeal swab collected using standard nylon flocked swab. Swab is inserted into the Viral transport medium (VTM) were provided from the same company (Molbio diagnostics Pvt. Ltd. Goa, India). Samples were transported immediately to the Molecular laboratory maintaining proper temperature and processed as per manufacturer’s guideline. [Truenat Beta CoV Chip-based Real time PCR test for Beta Coronavirus, Molbio diagnostics Pvt. Ltd. Goa, India]. The target sequence for this assay is E gene of Sarbeco virus and human RNaseP (serves as internal positive control). Confirmatory genes were RdRP gene and ORF1A gene.

### CMV detection

Whole blood was collected from the patients on 15 and 30 days of SARS-CoV-2 positivity, for detection of cytomegalovirus DNA. DNA extraction was carried out from whole blood using QIAamp® mini kit (QIAGEN GmbH, Hilden, Germany). Cytomegalovirus (CMV) detection and quantitation were carried out using CMV R –gene kit (bioMerieux SA, Marcy I’ Etoile-, France) by q-PCR on CFX96 Real time system (Bio-Rad, Hercules, CA). CMV viremia was defined as detection of greater than 5 × 10^2^ copies/ml.

### Immunological monitoring

Peripheral blood mononuclear cells (PBMC) were isolated from whole blood samples of the patients at days 15 and 30 following detection of SARS-CoV-2, by density gradient centrifugations using HiSep™ LSM 1077 media. For surface staining, 0.5 × 10^6^ cells were washed with phosphate-buffered saline (PBS) and stained with the following antibodies which were used for phenotypic analysis: CD3(APC-H7, SK-7) CD16 (PE-Cy7, B73.1), CD56 (APC R700, NCAM16.2), CD57 (BV605, NK-1), NKG2A (PE-Cy7, Z199), from BD Biosciences, (San Jose, CA) and NKG2C (PE, REA205) from Miltenyi Biotec, Germany. Cells were then incubated for 30 minutes. Viability was assessed with 7-AAD viability dye (Beckman Coulter). For intracellular staining, cells were stained for IFN-gamma using monoclonal antibodies for interferon-gamma (IFN-g) (4S.B3) and perforin (Alexa647, DG9) (BD Biosciences) after fixation and permeabilization with appropriate buffer (BD Biosciences and e-biosciences, San Diego, CA, USA). Flow Cytometry was performed using 10 colour flow cytometry (BD FACS Lyrics) and the flow cytometry data was analyzed using FlowJo software (v10.6.2, FlowJo). ANK cells were defined as CD56dimNKG2C+NKG2A-CD57+ subset of NK cells.

### KLRC2 (NKG2C) genotyping

KLRC2 gene encodes for NKG2C and is located in chromosome 12p13. DNA were isolated from the peripheral blood using QIAGEN QIAamp@ DNA blood mini kit method. PCR amplification was carried out with forward and reverse primer sequences as previously described(11) for wild type and deletion sequences of KLR2C gene (see supplement). The patients were categorised as KLRC2 wildtype homozygous (wt+/wt+), KLRC2 deletion homozygous (del+/del+) and KLRC2 deletion heterozygous (del+/wt+).

### Statistics

Total lymphocyte counts have been described, both in absolute numbers as well as % of total leucocyte count in Table 1. Subsequent lymphocyte subsets have been represented as % of the parent population. Binary variables were compared between the groups using chi square test. The continuous variables were analysed using independent sample t-test considering Levene’s test for equality of variances and non-parametric tests (Mann-Whitney U test). Probabilities of survival were estimated using the Kaplan-Meier product-limit method. P value < 0.05 was considered to be significant. Recursive partitioning analysis was carried out using *rpart* package (https://cran.r-project.org/web/packages/rpart/index.html) in R (https://cran.r-project.org/) to generate optimal cut off for ANK cells at day 30 in terms of outcome.

**Table 1:**
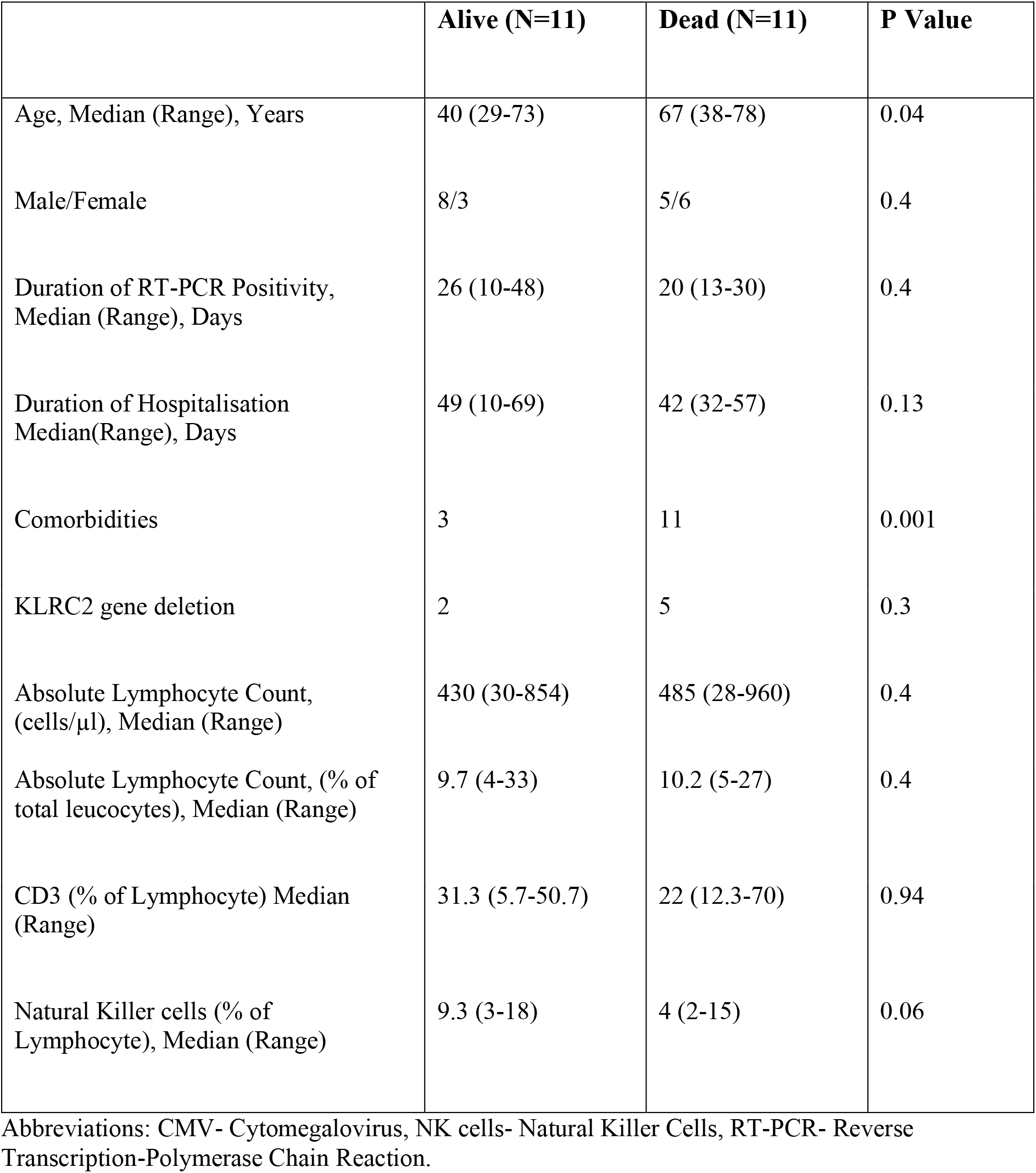
Patient Characteristics and Immunological parameters at day 30

|  | **Alive (N=11)** | **Dead (N=11)** | **P Value** |
| --- | --- | --- | --- |
| Age, Median (Range), Years | 40 (29-73) | 67 (38-78) | 0.04 |
| Male/Female | 8/3 | 5/6 | 0.4 |
| Duration of RT-PCR Positivity, Median (Range), Days | 26 (10-48) | 20 (13-30) | 0.4 |
| Duration of Hospitalisation Median(Range), Days | 49 (10-69) | 42 (32-57) | 0.13 |
| Comorbidities | 3 | 11 | 0.001 |
| KLRC2 gene deletion | 2 | 5 | 0.3 |
| Absolute Lymphocyte Count, (cells/μl), Median (Range) | 430 (30-854) | 485 (28-960) | 0.4 |
| Absolute Lymphocyte Count, (% of total leucocytes), Median (Range) | 9.7 (4-33) | 10.2 (5-27) | 0.4 |
| CD3 (% of Lymphocyte) Median (Range) | 31.3 (5.7-50.7) | 22 (12.3-70) | 0.94 |
| Natural Killer cells (% of Lymphocyte), Median (Range) | 9.3 (3-18) | 4 (2-15) | 0.06 |

Abbreviations: CMV- Cytomegalovirus, NK cells- Natural Killer Cells, RT-PCR- Reverse Transcription-Polymerase Chain Reaction.

## Results

Twenty-two patients with a median age of 56 years (range, 29-78) were evaluated (Table 1). All had severe bilateral pulmonary involvement as per CT-scan score(12) and received intensive respiratory support.

The median duration of RT-PCR positivity was 23 days (range,10-48). There was no relationship between mortality and persistence of positive RT-PCR. The median duration of illness (until discharge or death) was 47 days (range, 15-84). Co-morbidities were identified in 14 patients in the form of hypertension or diabetes mellitus or both. Eleven patients died of progressive respiratory failure at a median of 42 days (range 32-57). The overall survival in this cohort at a median of 60 days was 50% (95%CI, 39.3-60.7). The median age was higher in those with mortality (median-67 vs 40 years, p=0.04) and all had comorbidities (p=0.001) [Table1].

### CMV Viremia

All the patients were CMV seropositive. Only 2 patients had low level CMV viremia at 15 days (< 1× 10^3^ copies/ml). On day 30, 12 patients showed evidence of CMV viremia. There was no relationship with dose or duration of steroid therapy as this was similar for all patients in this study. Survival in patients with CMV viremia was 25% (95%CI, 12.5-37.5), compared to 80% (95%CI, 67.4-92.6) in those without viremia (p=0.01). The median CMV viral load was 1 × 10^3^ copies/ml (range, 0.6 × 10^3^ -135 × 10^3^). This was significantly higher for patients who died (median-22.4 × 10^3^/ ml vs 0 copies/ml in survivors, p=0.006, Figure 1).

**Fig.1.**
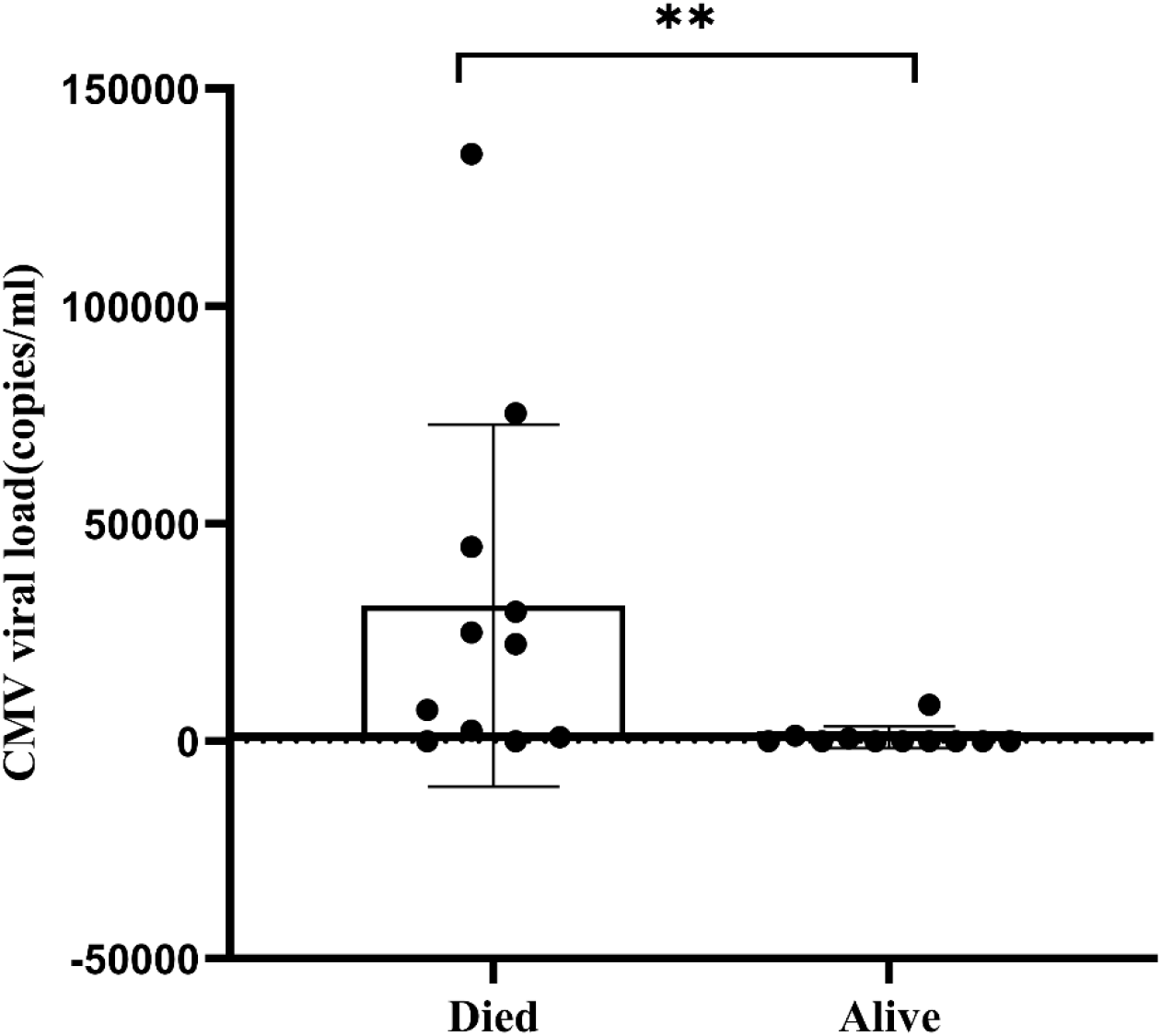
Scatter plot with bar showing difference in CMV viral load in terms of survival. **-p <0.01.

### Kinetics of Adaptive NK cells and Cytokine Release

Both absolute lymphocytes as well as CD3+ T cells were reduced in this cohort at 30 days. (Table 1). However, there was no impact of these parameters on mortality. NK cells were reduced as well in the overall cohort but did not correlate with mortality. ANK cells were markedly reduced in all patients (<5% of total NK cells) at day 15. However, at 30 days, this significantly improved in the survivors (14.1 ± 13%) and remained markedly reduced in those who died (2.2 ± 4.7 %), (p=0.0004, Figure 2). The inhibitory counterpart of ANK cells at 30 days, defined as NKG2A+/NKG2C-cells were significantly higher in those with mortality (83.1 ± 6.8 % vs 62.3 ± 20.4 %, p=0.002, Figure 2). Those with lower ANK cells tended to have greater CMV reactivation (p=0.07).

**Fig.2.**
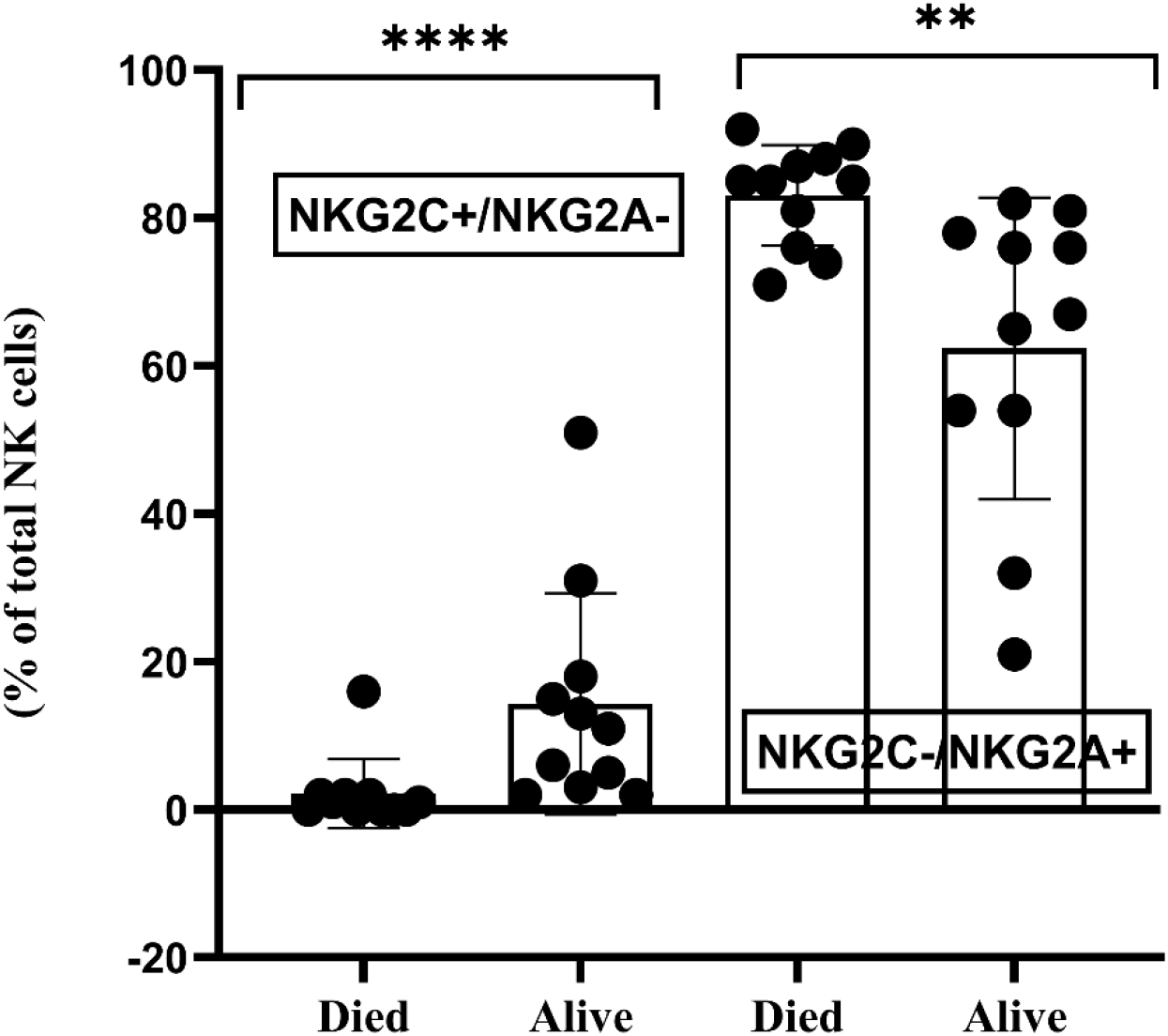
Scatter plot with bar showing NKG2C+/ NKG2A- and NKG2C-/NKG2A+ NK cells at Day 30 in patients in terms of survival. ***-p <0.001 and **-p <0.01

Cytokine release was evaluated at both days 15 and 30 on 5 patients each in survivors and non-survivors. Both IFN-g and Perforin release were markedly impaired at day 15 in all 10 patients. However, this improved significantly in the survivors, compared to those who died subsequently (Figure 3A-C, p < 0.01).

**Fig.3.**
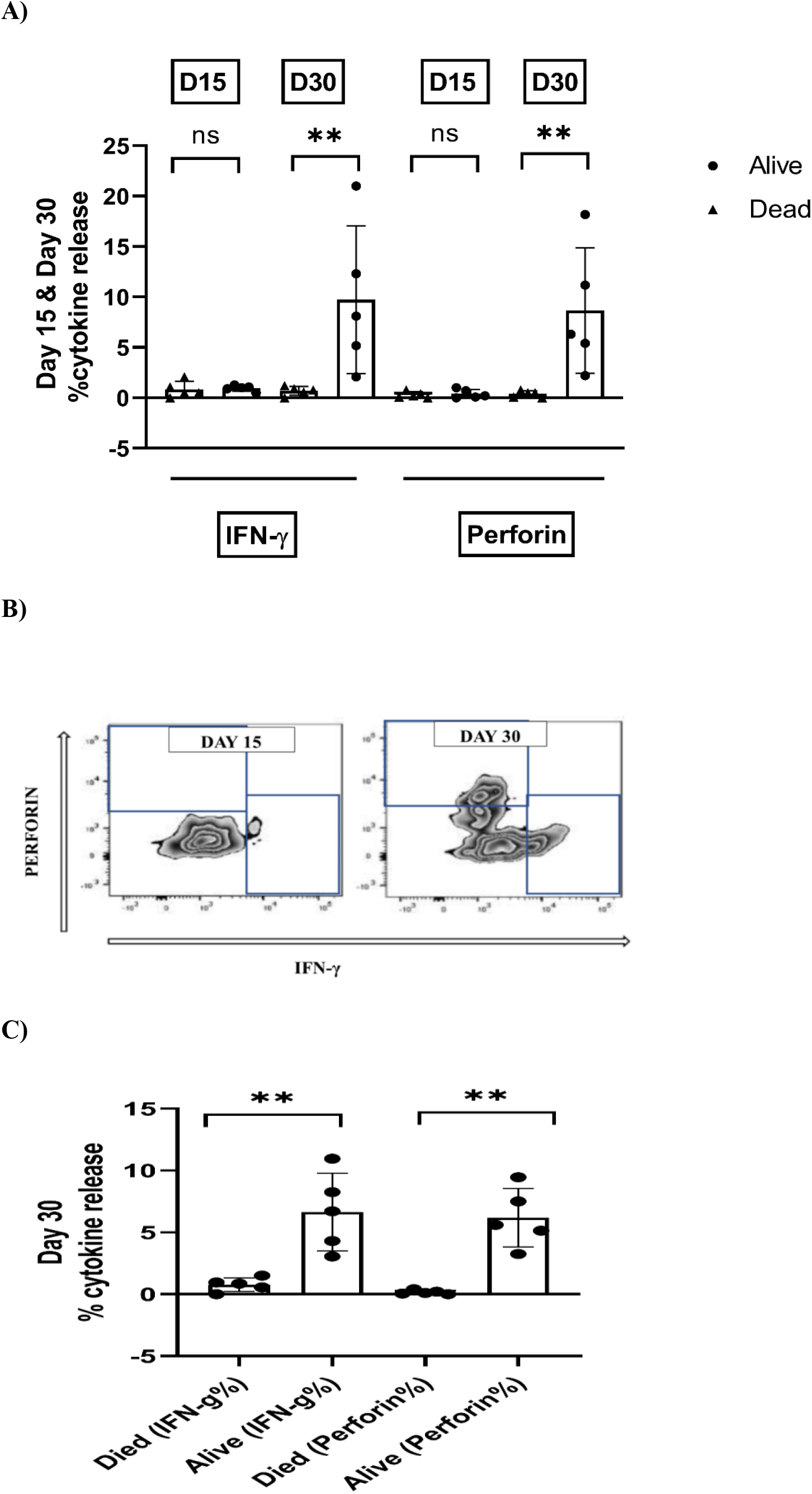
A. Symbols and lines plot showing difference in Cytokine release (IFN-g and perforin) between Day 15 and Day 30 in patients in terms of survival. **-p <0.01 and ns = not significant Fig.3. B. Flow cytometry plot showing interferon-g and perforin release in a survivor (UPN-6) at Day 15 and Day 30. Fig.3. C. Scatter plot with bar showing difference in IFN-g and perforin release at day 30 in patients in terms of survival. **-p <0.01

### KLRC2 deletion

Fifteen patients were KLRC2 wt+/wt+ and heterozygous KLRC2 deletion (wt+/del+) was detected in 7 patients. KLRC2 del genotype tended to be associated with CMV viremia (6/7 vs 6/15 without KLRC2 deletion, p=0.07). ANK cells were significantly lower in KLRC2 deletion group (2.2 ± 2.2%), compared to those with wild type genotype (10.96 ± 14.3%) [p=0.03, Figure 4]. Even though 5 out of 7 patients with KLRC2 deletion died, this was not statistically significant (p=0.3; Table 1).

**Fig.4.**
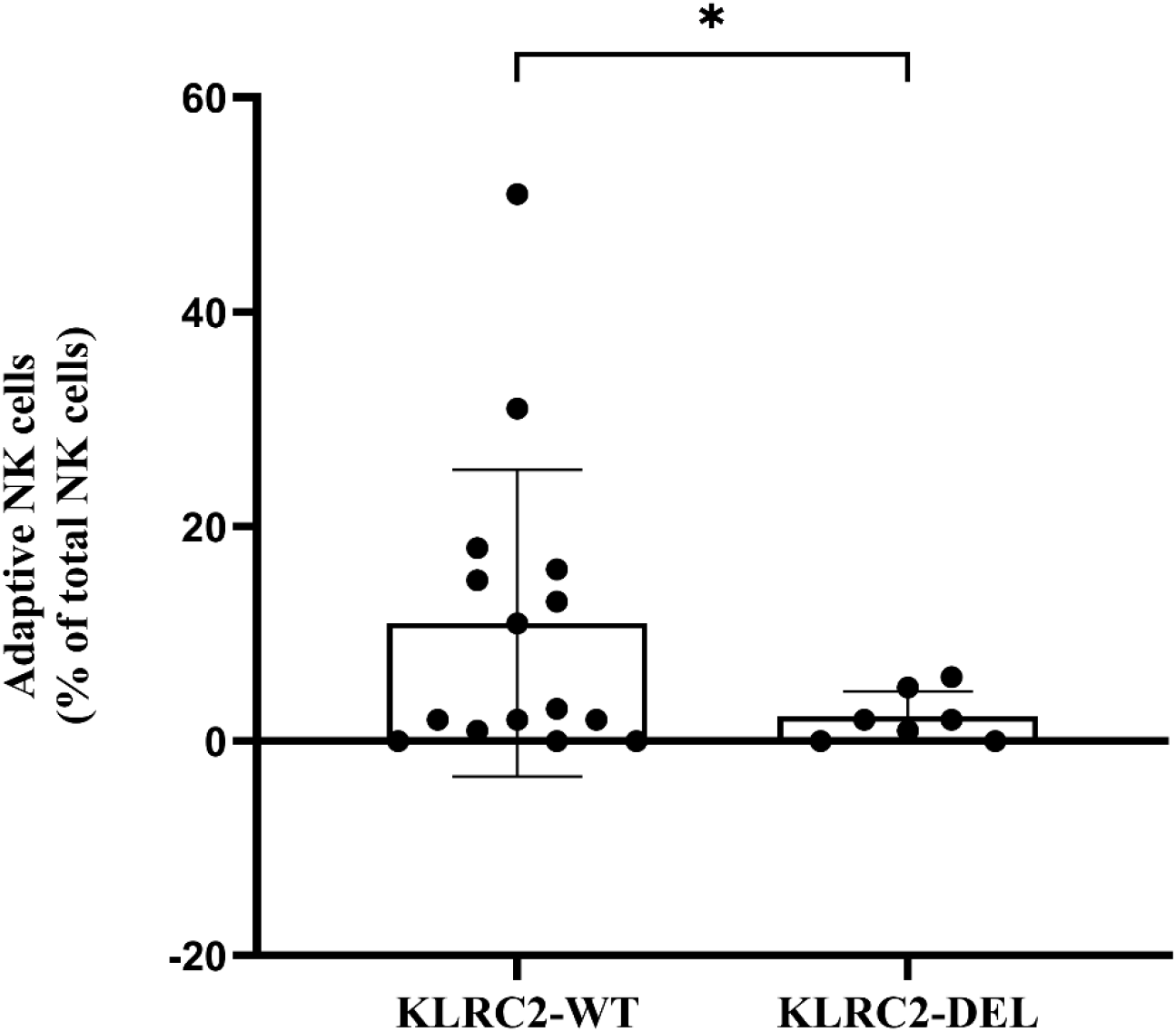
Scatter plot with bar showing difference in ANK cells at day 30 in patients with KLRC2 genotype, wildtype vs deletion, in terms of survival. *-p <0.05.

Based on recursive partitioning to determine the optimal cut point for absolute counts of ANK cells at day 30, survival was 90% (95%CI, 80.5-99.5) in those with ANK >2.5% vs 16.7% (95%CI, 5.9-27.5) in those below 2.5% (p=0.0001, Figure 5).

**Fig.5.**
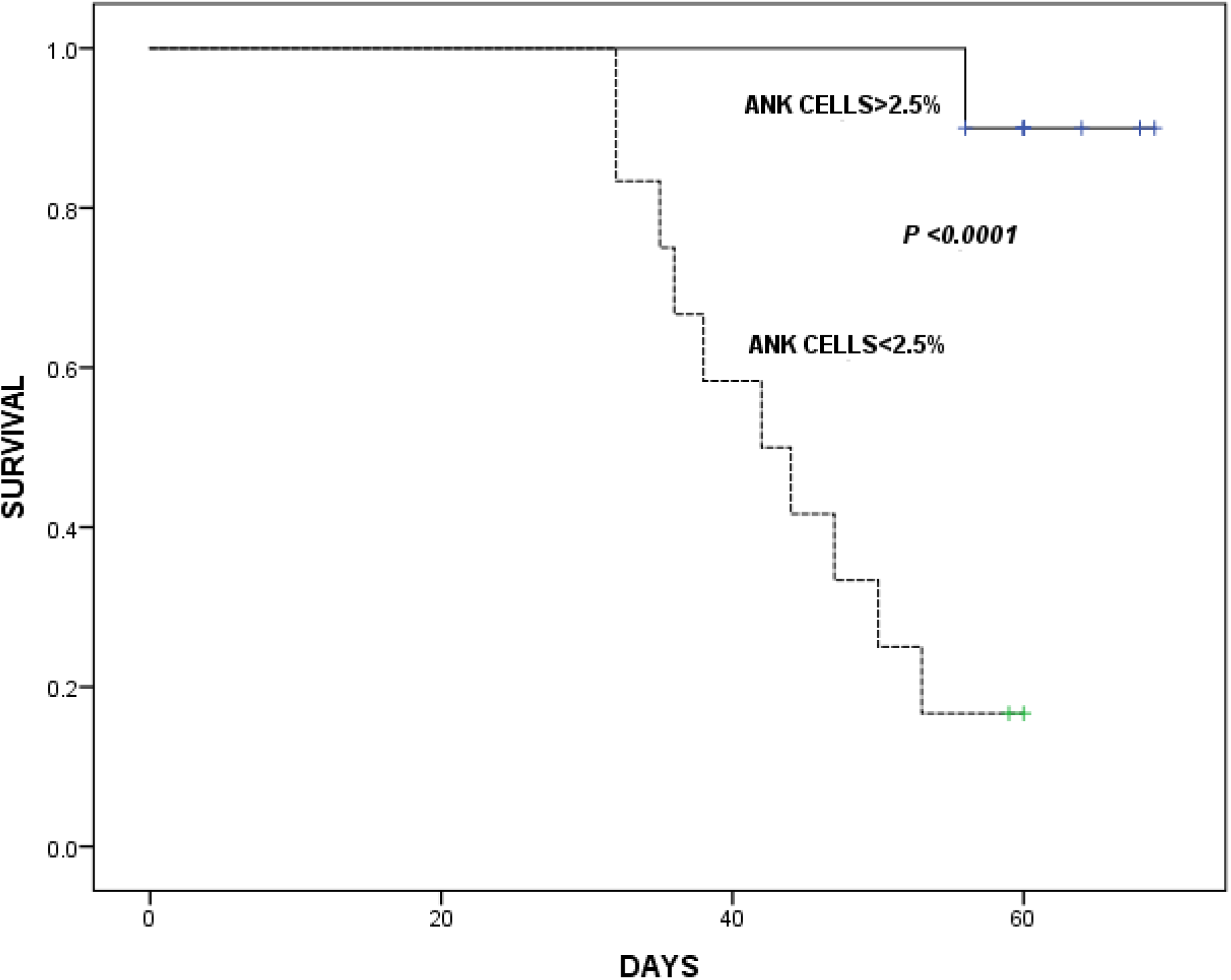
Kaplan-Meier plot showing survival in patients with Adaptive NK cells <2.5% (N=12) and >2.5% (N=10). ****-p <0.0001.

## Discussion

Immunology of COVID-19 has been at the centre of the discourse through the pandemic and NK cells have often been in the spotlight(13). Most of these studies have focussed on the NK cell profile in the acute phase in patients with differing disease severity. In that context, the current study addresses a cohort of patients with severe lung-disease, who survived the initial lung pathology beyond 30 days. Despite clearing the virus, over half the patients succumbed to progressive lung disease by 60 days. Our study focussed on the role of ANK cells in the etiopathogenesis of this phenomenon and the salutary findings in this small but unique cohort of patients highlight the role of ANK cells in CMV reactivation and subsequent adverse outcome.

While the contributory role of fungal pathogens in the late demise of COVID-19 patients have been well established(14), little data exist on the role of CMV reactivation(15-17). We found that majority of the patients had CMV reactivation, and this along with high CMV viremia were associated with higher mortality. In the setting of an allogeneic hematopoietic cell transplantation, high dose steroids have been strongly correlated with CMV infection and disease(18). It is important to note that although all patients received high dose steroids, there was no impact of the dose or duration of steroids on CMV reactivation. This brings to fore our endeavour to explore the impact of ANK cells on CMV infection and more importantly, how and if SARS-CoV-2 impacts the kinetics of ANK cells and vice-versa.

Very few studies have longitudinally studied the kinetics of ANK cells in the context of prolonged and severe COVID-19 disease. The studies so far have focussed almost exclusively on the acute phase of the disease with mixed reports. Several studies have emphasised on the high expression of NKG2A, a strong inhibitory counterpart of NKG2C, in those with severe disease(19). It has been demonstrated in an in-vitro study that spike protein-1 (SP1) of SARS-CoV-2 increases the expression of HLA-E on lung epithelial cells and NKG2A on NK cells as well, impeding both NK cell activation and viral clearance(20). On the other hand, a small study from Sweden suggested marked upregulation of ANK cells in the acute phase in those with severe COVID (9). However, the implications remain unclear due to the lack of corroborative data on outcome. Another study reported a correlation between KLR2C deletion genotype and disease severity without any data on NKG2C expression(10).

Our data showed a marked suppression of ANK cells in all patients in this cohort with severe disease at 15 days. There was significant upregulation of NKG2A+ NK cells. Cytokine release data showed marked functional impairment as well. Yet, a subset of patients showed both phenotypic as well functional recovery of ANK cells at 30 days, irrespective of clearance of SARS-CoV-2, which strongly correlated with a lack of CMV infection and a favourable outcome. On the other hand, those with persistently suppressed ANK cells, showing a high NKG2A phenotype, had CMV infection with high level of viremia and subsequent mortality. It is worth noting that majority of the latter group were older with comorbidities. Some researchers have suggested age-related CMV reactivation contributing to COVID-19 morbidity(21, 22). The only other study, which has systematically examined the incidence of CMV reactivation in patients with severe COVID-19 lung disease, detected pp65 antigenemia in 6 out of 26 critically ill patients, with a higher incidence of both bacterial and fungal infections in these patients, with a higher mortality. There was no impact of age, corticosteroid use or other inflammatory parameters on CMV infection. However, any immunological correlation of CMV reactivation in COVID-19 disease has been lacking to date and our study highlighting the strong correlation between ANK cells and subsequent CMV reactivation might help understand this phenomenon better.

While age or comorbidities could have possibly impacted the ANK cell recovery, a direct role of SARS-CoV-2 on downregulation of ANK cells cannot be discounted, given the in-vitro findings demonstrating the same(20). Either way, suppressed ANK population seemed to strongly correlate with CMV viremia and the final outcome. In the absence of autopsy studies, it is difficult to ascertain the exact contribution of CMV infection to the lung pathology. It is possible that CMV reactivation in this context might be a surrogate for severe immune suppression, particularly that of ANK cells, rather than being a primary cause for mortality.

One might speculate if the impact of ANK cells extend beyond its anti-viral effect. In this context, it is worth noting that inversion of NKG2C and NKG2A expressions following EBV reactivations, in the context of allogeneic HCT, strongly predisposed to chronic graft-versus-host disease(23). The same was demonstrated in the context of acute GVHD as well(7, 24). In contrast to conventional NK cells, which are suppressed by regulatory T cells (Tregs), ANK cells are inherently resistant to Treg-mediated suppression and vice-versa(25). Thus, ANK cells are uniquely placed to augment anti-viral cytotoxicity and downregulate exaggerated T cell reactivity at the same time.

Due to the small numbers in the cohort, no conclusive evidence could be drawn implicating KLRC2 genotype on the outcome, but the deletion phenotype was possibly contributory to the decremental ANK cells in the affected subset of patients. We have observed that KLRC2 wt+/del+ genotype does not necessarily correlate with compromised ANK cells in healthy adults and vice-versa. Thus, the impact of KLRC2 deletion genotype is unlikely to be independent of the kinetics of ANK cells and a heterozygous state is not necessarily associated with impaired NKG2C phenotype, unlike that suggested in a previous study extrapolating KLRC2 deletion status with low NKG2C expression(10). Our group is currently further exploring the highly variable levels of gene expression in those with KLRC2 heterozygous genotype, which might explain the variable levels of ANK cells in these individuals.

Thus, our study suggests that prolonged severe COVID-19 might have a detrimental impact on ANK cells, both phenotypically and functionally, which might persist in a subset of patients with higher age and comorbidities, contributing in turn to CMV infection and adverse outcome, even after viral clearance. Large prospective studies as well as further mechanistic explorations are needed to explore this further.

## Data Availability

All data generated in the present work are contained in the manuscript.

## Authorship Contribution

SRJ, AB and SC designed the study. JA, RL, MS and GB performed the study. SRJ, AS, and MS collected the data. SRJ, JA, AS and SC analyzed the data; SRJ and SC wrote the manuscript. All the co-authors reviewed and approved the manuscript.

## SUPPLEMENTARY FILE

### KLRC2 Genotyping by conventional PCR

DNA were isolated from the patients and donor samples from their peripheral blood using QIAGEN QIAamp@ DNA blood mini kit method (QIAGEN GmbH, Hilden, Germany). Isolated genomic DNA samples were used for the conventional PCR amplification. PCR amplification was carried out in a 20μl volume, containing 1x PCR master mix which has premixed taq polymerase, dNTPs, PCR buffer (Thermo fisher scientific, Waltham, MA 02451, United States), 1.65 pmol (for KLRC2 deletion) forward primer sequence: *5’ ACTCGGATTTCTATTTGATGC 3’* and reverse primer sequence: *5’ACAAGTGATGTATAAGAAAAAG 3’* and 1.65 pmol (for wild type KLRC2 gene) forward primer sequence: *5’ CAGTGTGGATCTTCAATG 3’* and reverse primer sequence: *5’ TTTAGTAATTGTGTGCATCCTA 3’* of each specific primer, 100pg-1μg genomic DNA. Amplification was performed using a T100 thermal cycler (Bio-Rad, Hercules, CA). Cycling temperature profiles were adopted from *Kai Cao et al*., with minor modifications. Briefly, the reaction mixture was subjected to one cycle of denaturation at 95^◦^C for 10 min followed immediately by 29 cycles of 95^◦^C for 20 s, 50^◦^C for 30 s, 72 ^◦^C for 40 s; and a final extension at 72^◦^C for 10 min before cooling to 4^◦^C. A non-template control for which double distilled water was used as DNA template, was included in each batch of PCR reactions. PCR products were identified by running entire PCR product on a 2% agarose gel for 60minutes at 70Volts. The size of the amplicons was determined by comparison against the migration of a 100-bp DNA ladder (GeneDireX, Inc, 333 taoyuan county, Taiwan). Agarose gel visualized and documented using the Gel doc XR+ gel documentation system (Bio-Rad, Hercules, CA).

## Supplementary Figure

**Supplementary figure 1:**
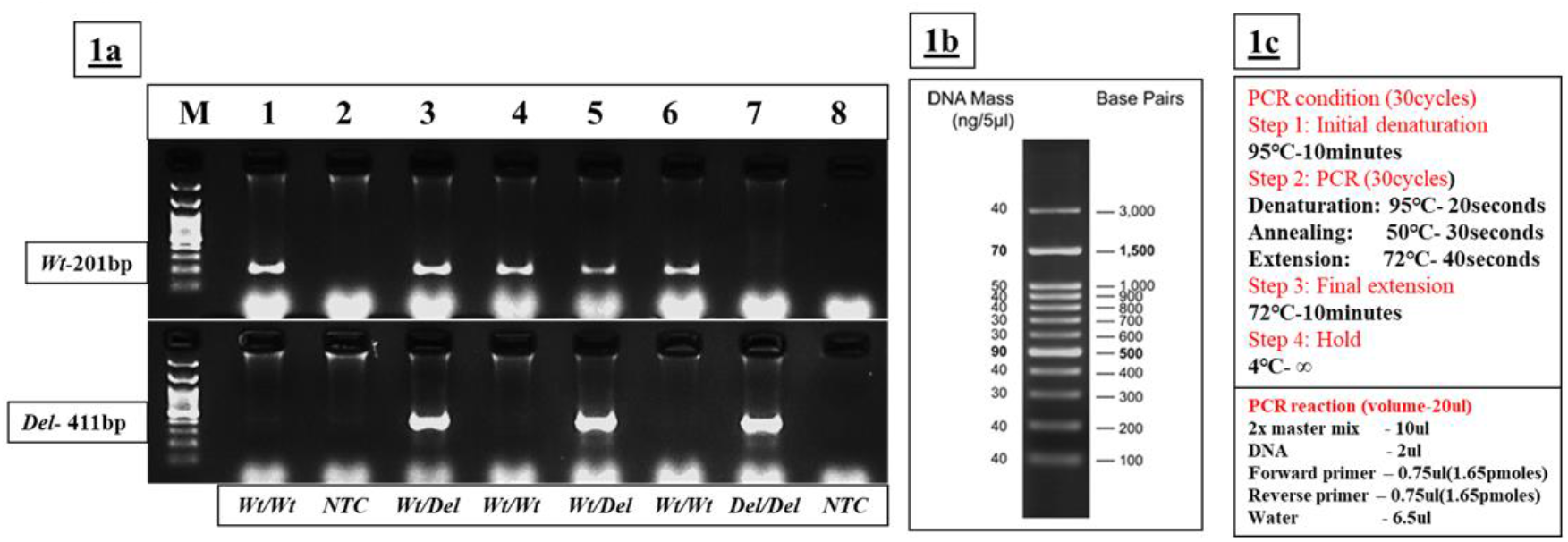
A)Above image represents the KLRC2 Conventional PCR gel image. DNA samples were isolated from the human samples. Isolated DNA samples were used for the **PCR amplification of KLRC2 Wild type(*Wt*) and Deletion (*Del)* gene (Primers details were mentioned in the methodology)**. For conventional PCR, 2µl of DNA samples were used and 30 cycles were used for amplification for both conditions. Top gel image shows the Wild type results and bottom image shows the Deletion type results. **Samples labelled as M-100bp DnA ladder, Samples 1-Homozygous Wild type positive control, Sample 2-NTC(non template control), Sample 3-Heterozygous both Wild type and Deletion positive control, Sample 4-Homozygous Wild type positive**, **Sample 5-Heterozygous both Wild type and Deletion positive, Sample 6-Homozygous Wild type positive, Sample 7-Homozygous Deletion positive Sample 8-NTC (Non Template Control). B)** 100bp DNA ladder data sheet image. **C)** PCR condition and PCR reaction preparation.

